# An ecological study of COVID-19 outcomes among Florida counties

**DOI:** 10.1101/2024.01.26.24301823

**Authors:** Sobur Ali, Taj Azarian

**Affiliations:** Burnett School of Biomedical Sciences, University of Central Florida, Orlando, Florida, USA

**Keywords:** COVID-19, Florida, Urban rural, Socioeconomic, Regression analysis, CFR

## Abstract

During the COVID-19 pandemic, Florida reported some of the highest cases and deaths in the US. County-level variation in COVID-19 outcomes has yet to be comprehensively investigated. Here, we assess corelates of COVID-19 outcomes among Florida counties that explain variation in case, mortality, and case fatality rates (CFR) across pandemic waves. County-level administrative data and COVID-19 case reports was obtained from public repositories. We tested spatial autocorrelation to assess geographic clustering of case rate, mortality rate, and CFR. Stepwise linear regression was employed to investigate the association between COVID-19 outcomes and 18 demographic, socioeconomic, and health-related county-level predictors. We found mortality rate and CFR were significantly higher in rural counties compared to urban counties, among which significant differences in vaccination coverage were also observed. Multivariate analysis found that the percentage of the population aged over 65 years, the percentage of obese people, and the percentage of the rural population were significant predictors of COVID-19 case rate. Median age, vaccination coverage, percentage of people who smoke, and percentage of the population with diabetes were significant influencing factors for CFR. Importantly, higher vaccination coverage was significantly associated with a reduction in case rate (R = -0.26, p = 0.03) and mortality (R = -0.51, p < 0.001). Last, we found that spatial dependencies play a role in explaining variations in COVID-19 CFR among Florida counties. Our findings emphasize the need for targeted, equitable public health strategies to reduce disparities and enhance population resilience during public health crises.

## Introduction

Severe acute respiratory syndrome coronavirus 2 (SARS-CoV-2), responsible for the pandemic of the coronavirus disease 2019 (COVID-19), has posed an unprecedented crisis to public health worldwide. As of December 2023, COVID-19 has resulted in 103 million cases and 1.14 million deaths in the US. As the pandemic unfolded, it became increasingly evident that demographic, socioeconomic, and health-related risk factors play crucial roles in influencing the COVID-19 outcome among communities in the United States (US) [1,2].

Socioeconomic predictors, including income, education, employment, and access to healthcare services, have long been recognized as critical determinants of health outcomes [3]. These factors may be associated with an individual’s capacity to adhere to public health guidelines, access healthcare resources, and implement preventive measures, which can impact the spread and severity of infectious diseases such as COVID-19. Furthermore, individuals residing in rural counties, characterized by socioeconomic challenges such as higher poverty rates, limited access to healthcare services, and lower educational attainment, have been found to be particularly vulnerable to COVID-19 [4,5]. However, variations in study time periods, population demographics, risk factors assessed, and endpoints have resulted in mixed findings among studies investigating the influence of demographic and socioeconomic risk factors on COVID-19 incidence rates [6].

Florida, one of the most populous states in the US, is notable for its unique demographic composition, high rates of immigration, recognition as a popular tourist destination, and climate. The diverse population, ranging from densely populated urban centers to sparsely populated rural counties, contributes to a complex web of potential influences on disease transmission and outcomes [7,8]. Considering the socioeconomic variation among counties in Florida, it is crucial to examine how these factors may have influenced COVID-19 outcomes.

In this study, we first investigated geographic clustering of COVID-19 outcomes. We then analyzed variation in case rate, mortality rate, and CFR across epidemic waves and assessed the association of demographic, socioeconomic, and health-related factors, and COVID-19 outcomes. Our results show that demographic, socioeconomic, and health-related differences explain a significant proportion of the variation in COVID-19 outcomes and that those inequalities disproportionately affected rural counties in Florida.

## Methods

### Map data

Florida Rural counties designations were obtained from the Florida Department of Health (FDOH) [9]. FDOH defines rural as (i) a county with a population of 75,000 or less, (ii) a county with a population of 125,000 or less which is contiguous to a county of 75,000 or less, (iii) any municipality within a county as described above [9]. County-level cartographic boundary Shapefiles were downloaded from the US Census Bureau TIGER Geodatabase [10].

### COVID-19 Outcome Data

County-level confirmed COVID-19 case and death data were obtained from The New York Times GitHub repository (https://github.com/nytimes/covid-19-data), based on reports from state and local health agencies [11]. Population statistics were obtained from the US Census Bureau (https://www2.census.gov/geo/docs/reference/ua/). Case rates represent the number of cumulative cases per 100,000 population and the mortality rate the number reported cumulated deaths from COVID-19 per 100,000 population as of December 2022. Each county’s CFR was determined by dividing the cumulative deaths associated with COVID-19 by cumulative COVID-19 cases [12].

We categorized the pandemic into five epidemic waves, delineated based on the epidemic curve and coinciding with the emerge of notable variants of concern during specific date ranges. The initial wave occurred from June 1, 2020, to September 30, 2020, corresponding to the onset of the pandemic and the original strain. Subsequent waves included the second wave from October 1, 2020 to May 30, 2021, associated with the Alpha variant; the third wave from July 1, 2021, to October 31, 2021, marked by the Delta variant; the fourth wave from December 1, 2021 to February 28, 2022, characterized by the emergence of the Omicron variant (mainly BA.1, BA.2 subvariant); and the fifth wave from April 1, 2022 to October 31, 2022, comprised largely of Omicron sub-variants BA.4 and BA.5.

### Covariates

Eighteen demographics, socioeconomic, and health-related factors were included in the study (**Table 1**). Data were collected from the Florida Community Health Assessment Resource Tool Set (CHARTS, www.flhealthcharts.gov ), maintained by FDOH. Data for the percent of the population in the county within rural blocks in Florida was obtained from the US Census Bureau and COVID-19 vaccination data for Florida counties from the Center for Disease Control and Prevention (CDC) [13]. Age-adjusted deaths from all causes per 100,000 population were collected from Florida CHARTS. Age-adjusted rates, which accounts to variation in county age structure, were calculated as described here (https://www.flhealthcharts.gov/charts/OpenPage.aspx?tn=665). All data used in this study were retrieved from publicly available databases.

**Table 1:** Definition and Summary statistics of different variables and COVID-19 outcomes in 67 countries in Florida. Mean, standard deviation (SD) and Coefficient of Variation (CV) of variables are listed.

| Variable | Description | Mean | SD | CV |
| --- | --- | --- | --- | --- |
| **Response Variables (COVID-19 outcomes)** |  |  |  |  |
| Case rate | Confirmed cases per 100,000 population | 30368 | 7163 | 23.59 |
| Mortality rate | Reported deaths per 100,000 population | 460.30 | 147.47 | 32.04 |
| Case fatality rate | Case Fatality Rate in percentage | 1.55 | 0.49 | 31.79 |
| **Demographic and socioeconomic factors** |  |  |  |  |
| Median age | Population median age | 44.03 | 6.54 | 14.85 |
| Aged | % of people aged over 65 years | 22.96 | 7.93 | 34.54 |
| Income | Median household income in Doller | 51290 | 10300 | 20.08 |
| Diploma | % Individuals with a high school diploma (aged 25 years and older) | 33.25 | 7.20 | 21.65 |
| Rural population | % of the population in the county within rural blocks | 37.50 | 32.26 | 86.02 |
| Unemployment rate | Unemployment rate, % of labor force | 6.64 | 1.53 | 23 |
| Poverty rate | % of total population, poverty rate | 15.18 | 5.10 | 33.6 |
| Below poverty level | % of total population, individuals below poverty level | 14.96 | 5.03 | 33.59 |
| **Health and behavioral factors** |  |  |  |  |
| Hospital beds | Hospital beds, rate per 100,000 population | 209.14 | 141.73 | 67.77 |
| Acute care beds | Acute care beds, rate per 100,000 population | 173.37 | 111.53 | 64.33 |
| Health Insurance | % of population, with any type of health care insurance coverage | 82.61 | 4.31 | 5.22 |
| Vaccinated one dose | % of population who received one dose of vaccine | 68.45 | 17.37 | 25.38 |
| Fully vaccinated | % of population fully vaccinated | 58.24 | 14.95 | 25.67 |
| Obesity | % of total population, who are obese | 32.46 | 6.07 | 18.69 |
| Diabetes | % of total population, who have ever been told they had diabetes | 13.37 | 3.09 | 23.11 |
| Smoking | % of total population, who are current smokers | 19.14 | 5.08 | 26.55 |
| **Race and Ethnicity** |  |  |  |  |
| Black | % of Population Black (of total population) | 14.86 | 9.38 | 48.24 |
| Hispanic | % of Population Hispanic (of total population) | 14.59 | 13.09 | 89.75 |

### Statistical Analysis

All variables were reported as mean and standard deviation, and the coefficient of variation (CV) was calculated to assess the relative dispersion. Spread and collinearity of the predictor variables were assessed through histograms, bivariate scatterplots, and Spearman correlation coefficients using R-package psych v2.3.6 [14]. Correlation coefficients were calculated for each predictor with the outcome using Spearman or Pearson correlation according to data type and distribution. Florida county-level case rates, mortality rates, and CFR were visualized in the map using ggplot2 v3.4.3 and sf v1.0.14 package in R v4.3.1.

We used multivariate ordinary least squares (OLS) regression model to examine the relationship of predictor variables and COVID-19 outcomes. Then, we built multivariate models for each outcome using a stepwise backward procedure, starting with the fully saturated model, and then iteratively dropping variables. Akaike Information Criteria (AIC) and Variance Inflation Factor (VIF) were used to assess model fit and the degree of collinearity among covariates, respectively. R^2^ was calculated for the final model [15]. All statistical analysis and visualization were performed using R v4.3.1 [16] with Rstudio v2023.06.1 [17] statistical software.

### Spatial autocorrelation

We calculated the global Moran I statistic to examine whether county COVID-19 case rate, mortality rate, and CFR were spatially autocorrelated [18]. First-order Queen contiguity binary neighbor list and spatial weights matrix were constructed using R package spdep v1.2.8. Counties were considered neighbors when they shared one common point or vertex [19]. Local spatial autocorrelation identified four distinctive categories: High-High, Low-Low, High-Low, and Low-High. Local spatial clusters of COVID-19 outcome were measured using GeoDa v1.22 software [20].

We compared the final selected ordinary least squares (OLS) model and two spatial regression models: spatial lag model (SLM) and spatial error model (SEM). SLM examines how the adjacent counties influence an index county’s outcome variable. The spatial lag parameter (r) estimates the association between the average logged period rate in nearby counties and the logged period rate in the index county. In SEM, the parameter (l) determines how well a county’s ordinary least squares (OLS) residual correlates with the residuals of its neighbors. This spatial error parameter (l) represents the strength of the association between the average residuals or errors in nearby counties and the residual or error of the current county [21]. We used the R “lagsarlm” and “errorsarlm” functions from spatialreg package to perform spatial regression analysis [22].

## Results

### Descriptive mapping of COVID-19 outcomes in Florida

Florida comprises 67 counties (**Figure 1A**) with a population of 21 million, where a higher percentage of the state’s population (87.7%) lives in urbanized areas. By December 2022, Florida reported around seven million confirmed cases and around 84,000 COVID-19 associated deaths. Visualization of cumulative rates across the five waves showed geographical heterogenicity in COVID-19 case rate, mortality rate, and CFR across Florida counties. Seminole, Miami-Dade, and Broward Counties had the highest case rate, while Glades, St. Johns, and Sumter Counties exhibited lower rates (**Figure 1B**). Union, Suwannee, Citrus Counties had the highest death rate, whereas Orange, Monroe, and St. Johns Counties had the lowest mortality rate (**Figure 1C**). Citrus, Highlands, and Union counties had the highest CFR and Leon, Orange, Monroe counties had lowest CFR (**Figure 1D**).

**Figure 1:**
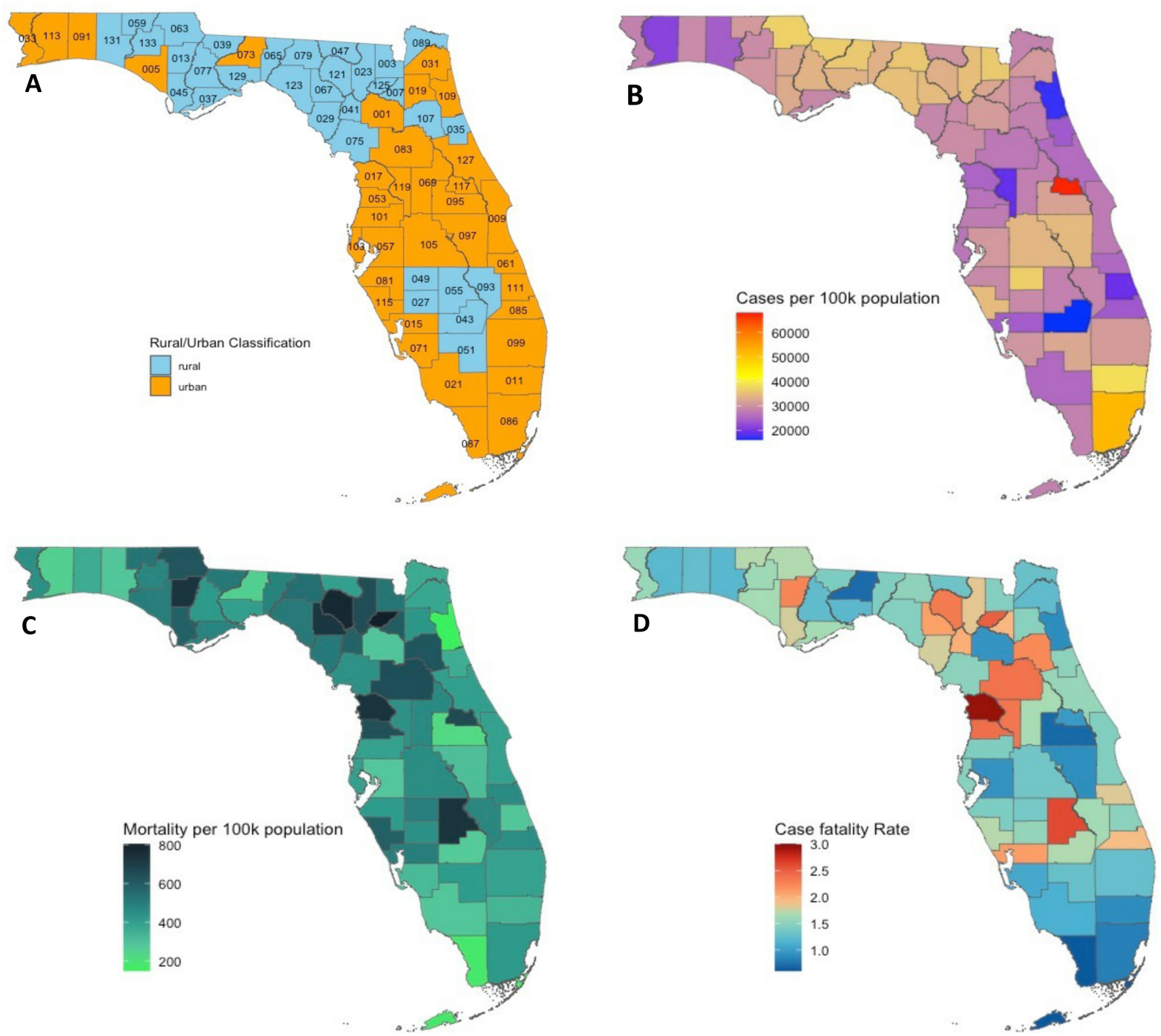
(A) Florida map showing geographic distribution of urban and rural counties. Statewide distribution of county-level COVID-19 confirmed cases per 100,000 population (B), mortality per 100,000 population (C), case fatality rate (D) in Florida from March 2020 to December 2022.

**Table 1** presents the summary and descriptive statistics of COVID-19 outcome variables and independent variables, and CV, which informs dispersion of the data. Mortality rate and CFR were more dispersed than case incidence rate. Demographic, socioeconomic, and health-related risk factors vary widely among counties. The percentage of people in the rural areas, hospital bed rate, Hispanic population, and poverty rate are the most heterogeneous factors.

### Spatial autocorrelation of COVID-19 outcome between Florida counties

Moran’s I statistics were calculated to variables among Florida counties. Assessment of spatial autocorrelation in COVID-19 outcomes indicated significant but weak positive spatial autocorrelation for case rate (I = 0.12, p = 0.037), mortality Rate (I = 0.22, p =0.001), and CFR (I = 0.3, p < 0.00). The Local Moran test was conducted to examine spatial clustering in more detail. The “High-High” category signifies areas with high values neighboring other counties with high values, “High-Low” counties with high values neighboring counties with low values, and “Low-Low” counties with low values neighboring other counties with low values. Orange County (Orlando, FL) belonged to a High-High case rate cluster, and Flagler, Putnam, Marion, Citrus and Hernando County a Low-Low cluster (**Figure 2A**). Madison, Suwannee, Lafayette, Gilchrist, and Columbia Counties belonged to a High-High mortality rate cluster, and Okaloosa, Collier, and Miami-Dade Counties a Low-Low cluster (**Figure 2B**). For CFR, we identified one High-High cluster consisting of Marion, Citrus, Sumter, and Hernando Counties and a Low-Low cluster consisting of four additional counties (**Figure 2C**).

**Figure 2:**
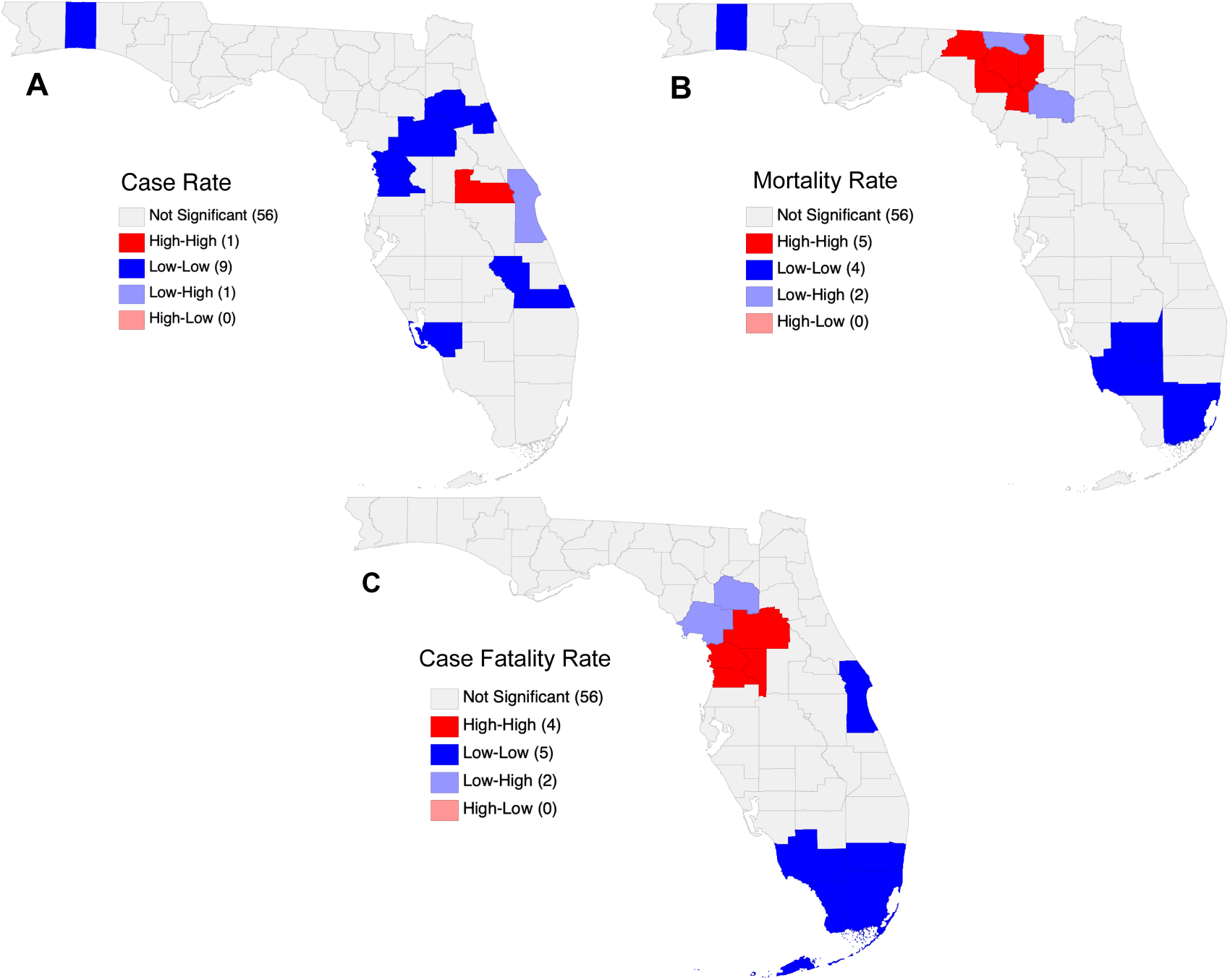
Spatial autocorrelation of COVID-19 outcomes by Florida county considering queen contiguity weights. Significant spatial clustering for case rate (B), mortality rate (C), and CFR (D) was observed. The High-High cluster (red) indicates counties with high values of a variable that are significantly surrounded by regions with similarly high values. The “Low-Low” cluster (blue) refers to counties where low COVID-19 outcome is surrounded by other counties with low COVID-19 outcome.

### Trends in COVID-19 case and mortality rates

Our analysis revealed higher case rates in the first and third waves in rural counties compared to urban counties. However, no significant difference in case rates was observed for the second and fourth waves (**Figure 3A**). Furthermore, we observed a noteworthy temporal difference. Over the five pandemic waves, rural counties appeared to experience a more acute increase in COVID-19 case incidence, while urban areas described by a more protracted epidemic curve (**Figure 4**). Our analysis indicates that, despite the differences in epidemic trajectories, cumulative COVID-19 case rates did not differ significantly between urban and rural counties (**Figure 3B**).

**Figure 3:**
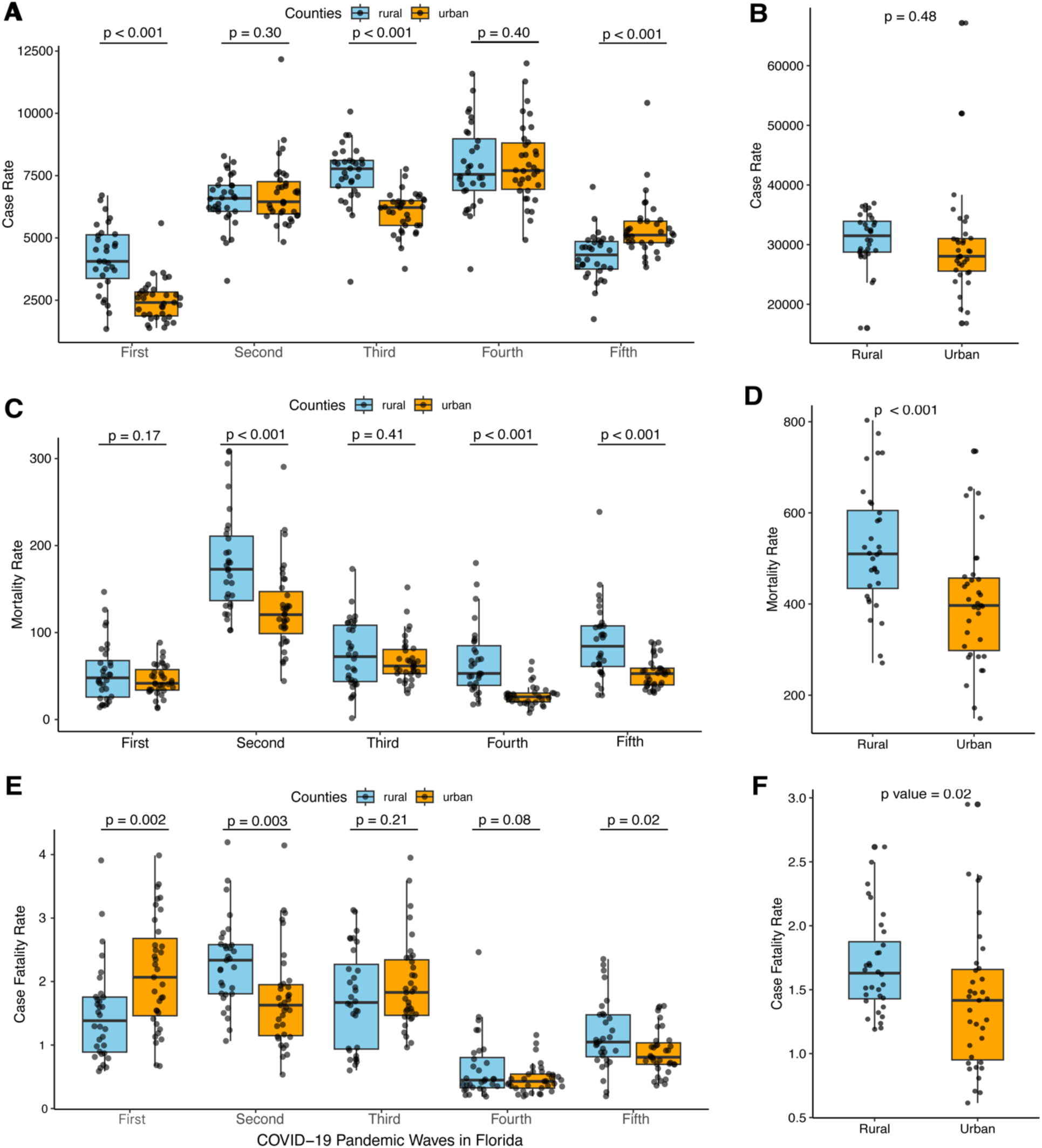
Variation in COVID-19 outcomes in Florida. (A) Variation of case rate in urban and rural counties in five pandemic waves and (B) difference in cumulative case rate (B) in urban and rural counties. (C) Variation of mortality rate in urban and rural counties in five pandemic waves and (D) difference in cumulative mortality rate. (E) Difference in CFR in urban and rural counties in five pandemic waves and (F) difference in overall CFR in Florida. Significance was assessed using a t-test. Data distributions are presented as box plots. Each dot point indicates the data of each county.

**Figure 4:**
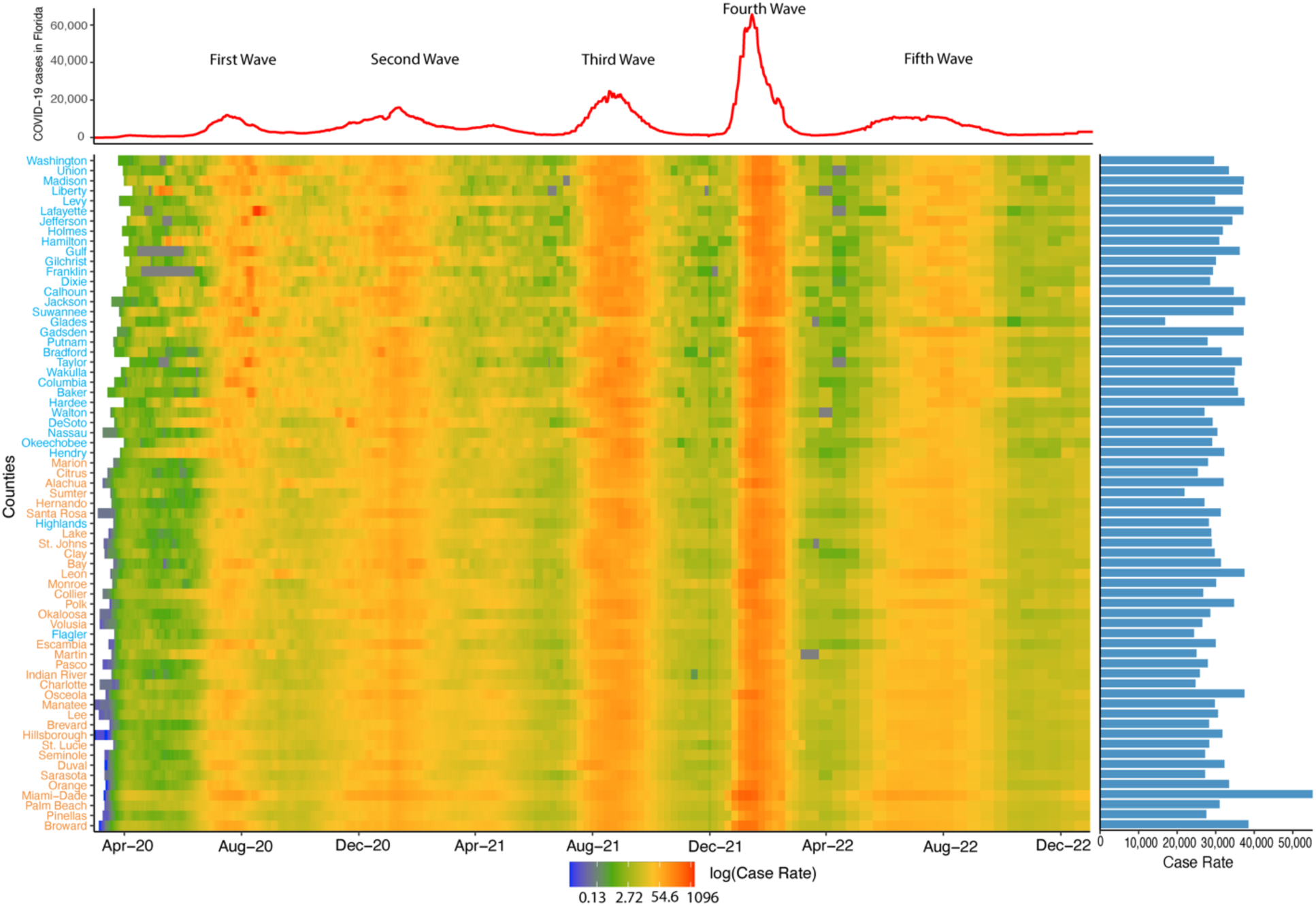
Trend in COVID-19 case rate among Florida counties. The heat map illustrates daily COVID-19 cases per 100,000 population with red coloration indicating higher values, and blue coloration indicating lower values. Counties are sorted by rural population percentage, with higher percentages at the top and lower percentages at the bottom. County names are color-coded, with orange indicating urban counties and blue representing rural counties, as defined by the Florida Health Department. The bar plot on the right side of the figure shows each county’s COVID-19 case rate per 100,000 population from the pandemic’s beginning to December 2022. At the top, a trend line displays the 7-day moving average of daily COVID-19 cases in Florida, with annotations marking five significant pandemic waves as of December 2022.

We further assessed mortality rates in urban and rural counties across five pandemic waves in the context of SARS-CoV-2 variants (**Figure 5**). We found that the mortality rates in rural counties was significantly higher than in urban counties during the second, third, and fifth waves (**Figure 3C**). While the difference in mortality rate between urban and rural counties during the first and fourth waves was not significant, the overall mortality rate was significantly higher among rural counties (**Figure 3D**). Variation in the case and mortality rate trends across the five waves of the pandemic was also observed. In particular, the third wave (Delta variant) exhibited the highest mortality rate, while the fourth wave (Omicron variant) showed a significantly higher case rate.

**Figure 5:**
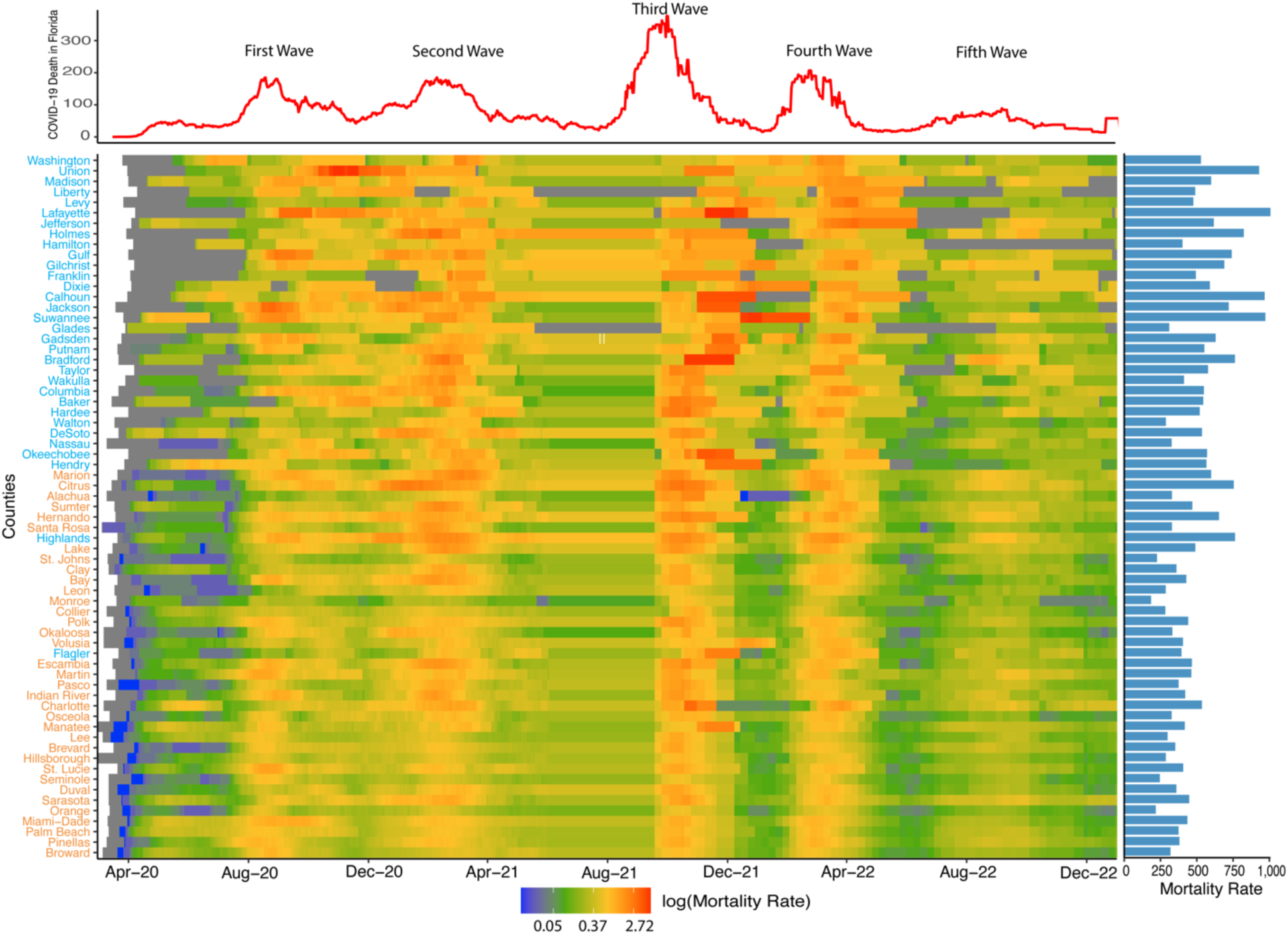
Trend in COVID-19 mortality rates among Florida counties. The heat map illustrates daily COVID-19 deaths per 100,000 population with red coloration indicating higher values, and blue coloration indicating lower values. Counties are sorted by population percentage within rural blocks, with the county having the highest percentage at the top. County names are color-coded, with orange indicating urban counties and sky blue representing rural counties, as defined by the Florida Health Department. At the top, a trend line displays the 7-day moving average of daily COVID-19 associated deaths in Florida, with annotations marking five significant pandemic waves. The right side presents the county-wise mortality rate as of December 2022.

CFR was significantly higher (p = 0.02) in rural counties than in urban (**Figure 3F**). However, we observed inconsistent patterns among urban and rural counties when we assessed CFR differences during the five waves. In the first wave, CFR was significantly higher in urban counties, whereas in rural counties CFR was higher in the second and fifth waves (**Figure 3E**). As decreased access to SARS-CoV-2 testing among rural counties may have resulted in an under estimation of case rates, we conducted a sensitivity analysis finding that even after accounting for a 2.6% underreporting of cases among rural counties, the CFR remained significantly higher (p = 0.04).

Lastly, we extended our investigation to include age-adjusted all-cause mortality rates, comparing urban and rural Florida counties before and during the pandemic. We observed higher all-cause mortality among rural counties than urban counties before and during the pandemic (**Figure 6A**). Our analysis also indicated that the difference in death rates between urban and rural counties increased during the pandemic (**Figure 6A**). By calculating the percent change in the age-adjusted all-cause death rate from 2019 (pre-pandemic) to subsequent pandemic years (2020, 2021 and 2022), we found a consistent and statistically significant trend: rural counties experienced a higher mean percentage change in age-adjusted all-cause mortality than urban counties across the analyzed years (**Figure 6B**).

**Figure 6:**
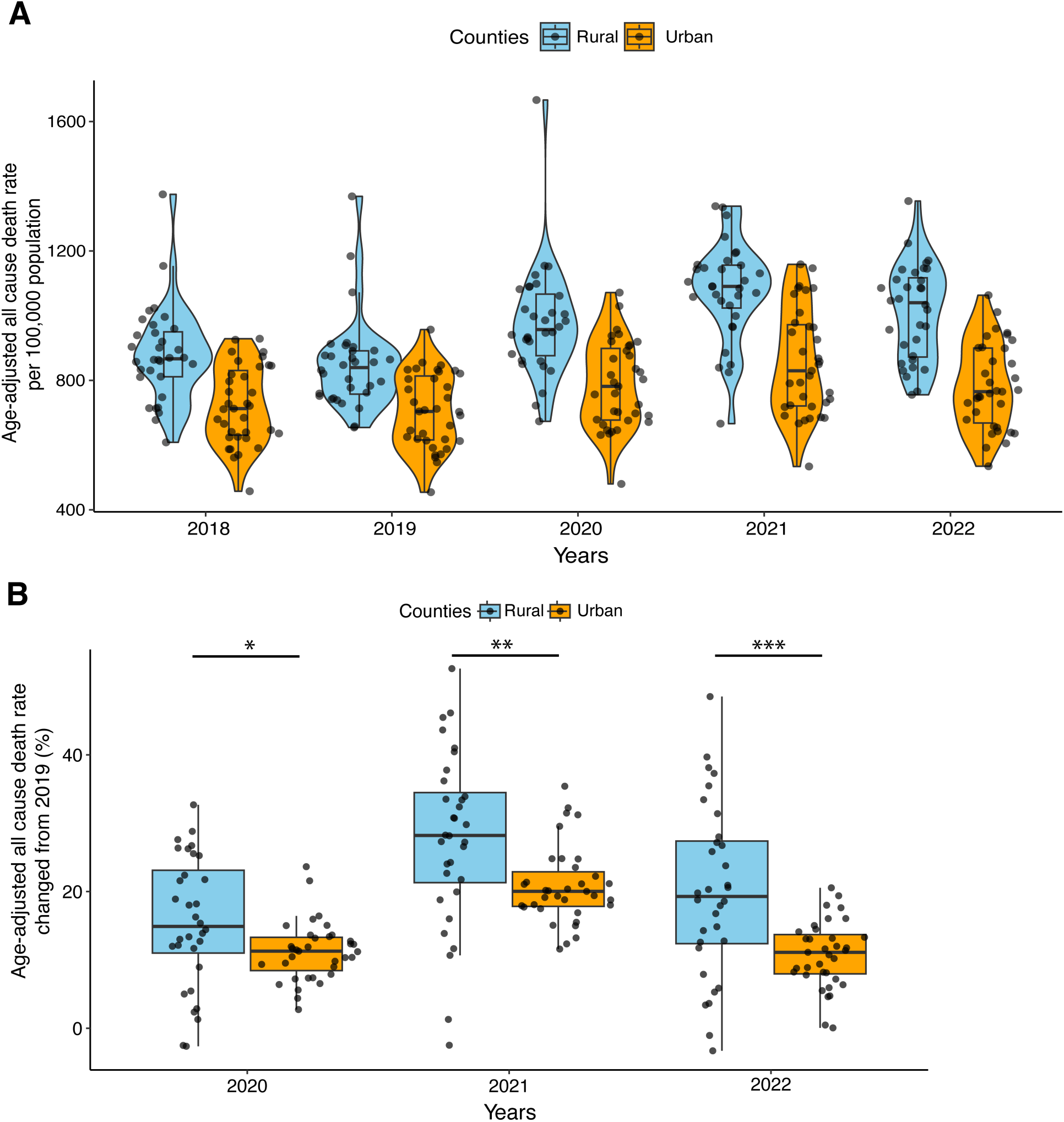
Age-adjusted all-cause death rate (per 100k population) in Florida during pre-pandemic and pandemic times. (A) Violin plots and boxplots of the age-adjusted all-causes death rates in urban (orange) and rural (sky-blue) counties in pre-pandemic (2018-19) and pandemic years (2020-22). (B) Boxplot presents the age-adjusted all-cause death rate change from pre-pandemic (2019) to pandemic times (2020-22) in urban and rural counties. For the t-test between urban and rural counties, significant differences are represented with asterisks (∗, p < 0.05; ∗∗, p < 0.01; ∗∗∗, p < 0.001).

### Vaccination disparities between urban and rural counties in Florida

To investigate the role of vaccination on the trajectory of the pandemic, we analyzed differences in vaccination coverage, finding significantly lower vaccination coverage in rural counties compared to urban counties (**Figure 7A**). Further, when considering vaccination trends, a persistent pattern became evident. Urban counties (Mean: 69.48, 95% CI: 66.55-72.41) consistently had ∼ 20% higher vaccination coverage than rural counties (Mean: 45.94, 95% CI: 42.39-49.50) (**Figure 7B**). However, several rural counties were outliers, exhibiting higher vaccination rates than certain urban counties.

**Figure 7:**
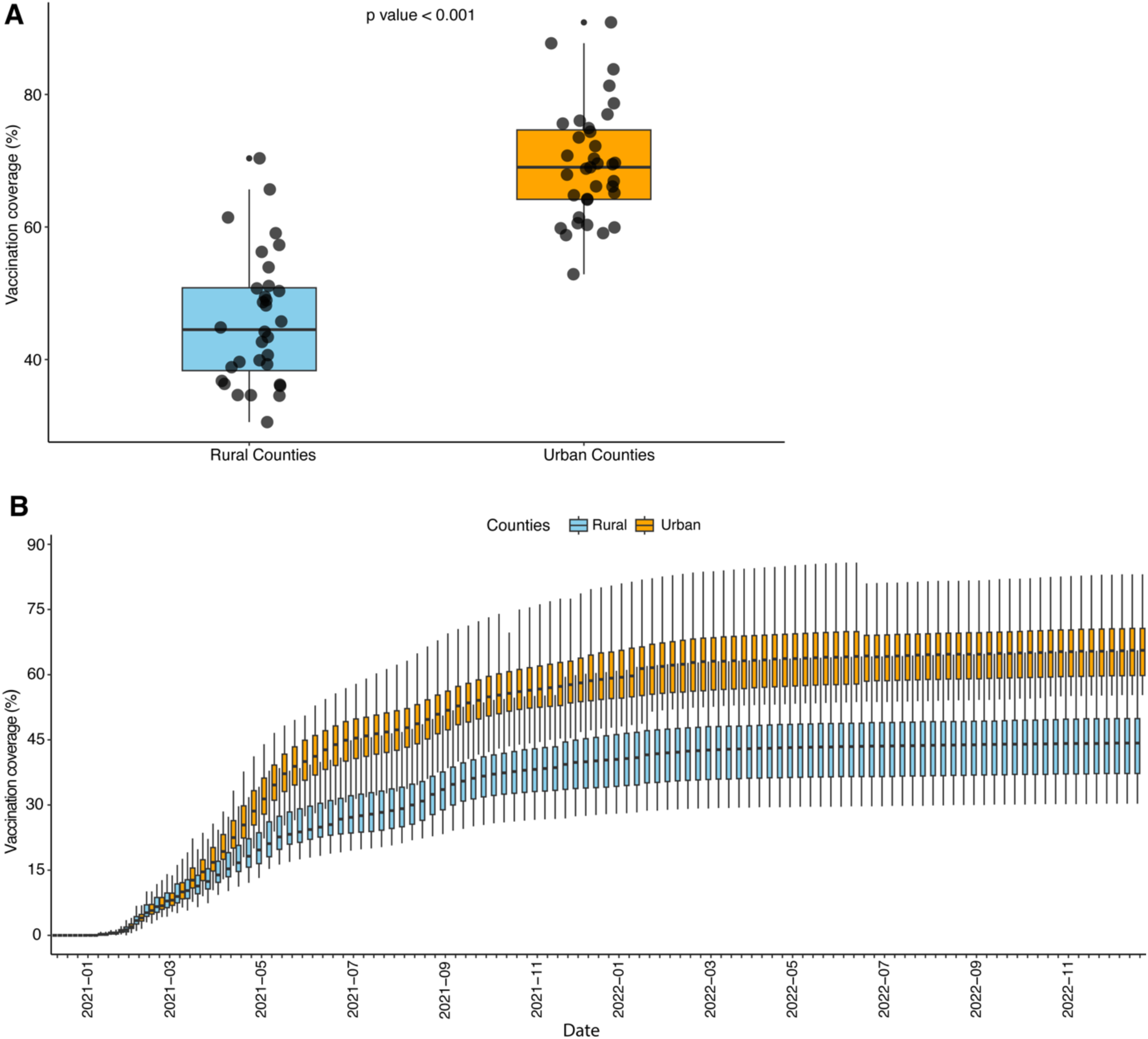
The variation of vaccination coverage in Florida. (A) Boxplot shows the vaccination coverage in urban counties(orange) and rural counties (sky-blue) in Florida. (B) The trend of COVID-19 vaccination coverage in urban and rural counties from December 2020 to December 2022.

### Correlation analysis

We evaluated the correlation between selected influencing factors and COVID-19 case rate, mortality rate, and CFR. A significant positive correlation was observed between case rate and individuals below poverty (r = 0.36, P = 0.003) and percent of the population of black race (r = 55, P < 0.001); significant negative correlation was observed between case rate and median age (r = -0.58, P < 0.001), percent of population insured (r = -0.32, p=0.008) and COVID-19 vaccination coverage (r = -0.26, P = 0.03). We also found a significant positive correlation between mortality rate and percent of rural population (r = 0.53, P = <0.001), percent of population with obesity (BMI>30) (r = 0.34, P = 0.006), percent of population with diabetes (r = 0.43, P < 0.001), and percent of population who are smokers (r = 0.48, P < 0.001). In contrast, a significant negative correlation was observed between the mortality rate and median household income (r = -0.52, P <0.001) and vaccination coverage (r = -0.51, P < 0.001). The CFR is significantly correlated with median age (r = 0.43, P <0.001), rural population (r = 0.41, P <0.001), diabetes (r = 0.50, P <0.001), and smoking (r = 0.39, P = 0.001) (**Table 2**).

**Table 2:**
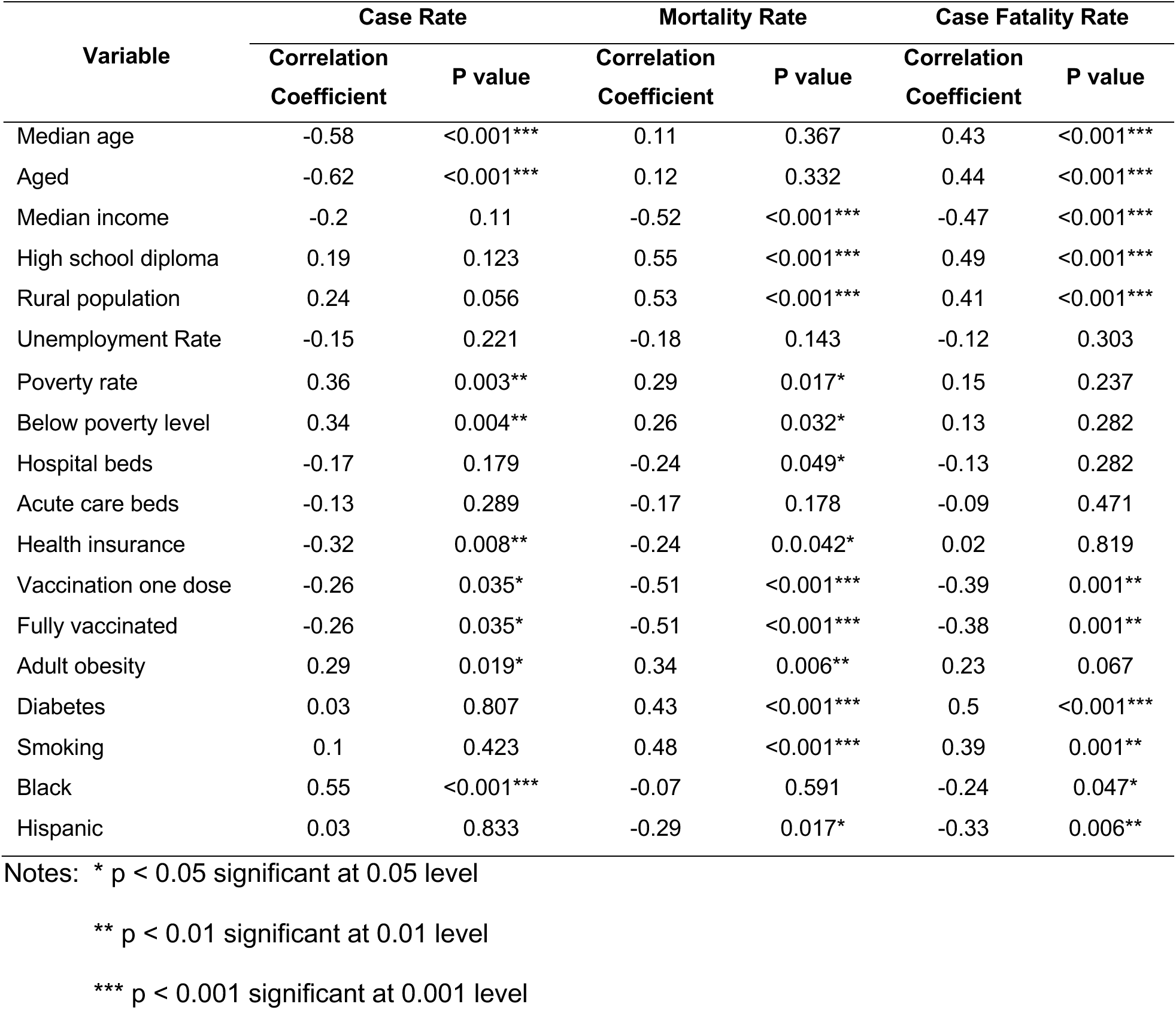
Univariate correlation analysis with predictor variables for case rate, mortality rate, and CFR.

### Multivariate analysis

The spread and collinearity of covariates were assessed (**Supplementary Figure S1**). COVID-19 vaccination with one dose and fully vaccinated was positively correlated, and as a result, we retained fully vaccinated in the final analysis. For all other highly correlated variables, such as hospital and acute care beds, which are both proxies for healthcare access, we retained only one variable for the multivariate analysis.

In the final model for case rate, the percentage of the population in counties with rural blocks, percent of black population, percent of obese population, Hispanic population, and percent of people aged over 65 years remained in the final stepwise model (R^2^=0.42, P < 0.01). A low value of VIF (< 3) suggested that the covariates in the final model did not have significant multi-collinearity and have substantial power to explain maximum model variation (**Table 3**).

**Table 3:**
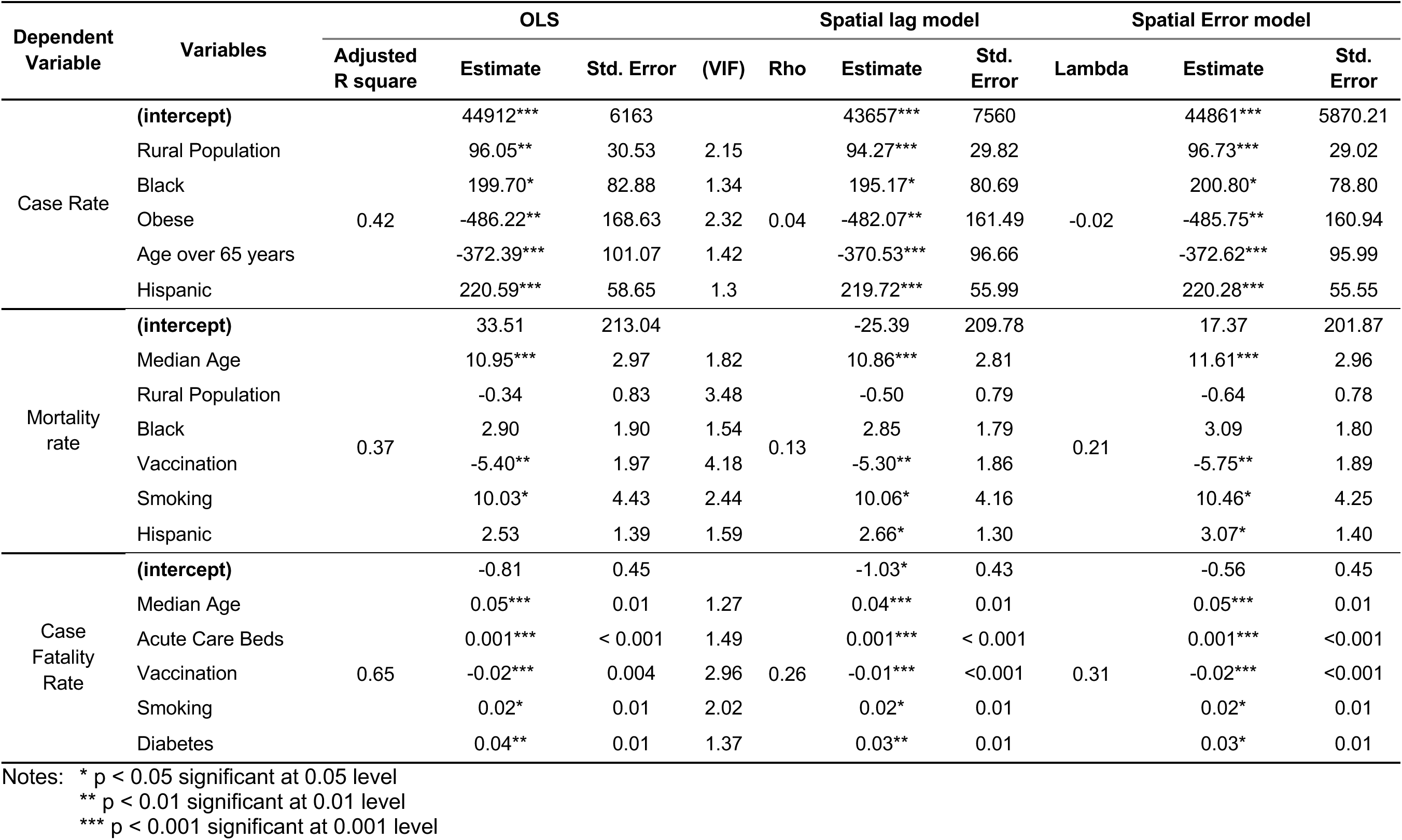
Multivariable regression models of the association between socioeconomic characteristics and risk factors with the COVID-19 outcome variables. The three outcome variables included are case rate, death rate, and case fatality rate. All are calculated per 100,000 people, and data was included up to December 2022.

For the mortality rate, the final selected model included median age, percent of people in the rural area, percent of black population, vaccination coverage, percent of population who smokes, and percentage of Hispanic population, which explained 37% of variation of mortality rate in Florida. The result of collinearity diagnostics (VIF < 5) indicated that the collinearity between predictor variables was acceptable in this multiple linear regression model (**Table 3**).

Based on the stepwise multiple linear regression analysis, median age, acute care beds rate vaccination coverage, percent of the population who smokes, percent of the diabetes population were found to explain 65% of variation in the CFR in Florida (**Table 3**). The collinearity diagnostics (VIF < 3) suggested that the predictor variables are relatively independent of each other in the final model.

### Spatial regression

We compared the final OLS models for case rate, mortality rate, and CFR with SLM and SEM to assess spatial association between the predictor covariates and outcome variables. For the case rate and mortality rate, coefficients of the OLS and SEM models are closer than SLM (**Table 3**). However, AIC values were lowest for the OLS model compared to the spatial models, indicating that case and mortality rate can be explained better by OLS (**Table 4**). We then explored SLM and SEM models to account for spatial dependencies in CFR. Notably, the estimates and significance of the covariates in the SLM model remained consistent with the OLS model, indicating that the spatial lag effects did not significantly alter the relationships between predictors and CFR. The SLM also estimated a spatial lag coefficient (rho) of 0.26, informing the extent to which neighboring counties’ CFR values influence each other. As with SLM models, the predictor variables of SEM exhibit estimates and significance similar to the OLS model. Even after accounting for these predictor variables, the SEM estimated a spatial error coefficient (lambda) of 0.31, indicating persistent spatial variation in CFR (**Table 3**). The AIC values further informed our model selection, with the SLM having the lowest AIC, indicating the best trade-off between model fit and complexity. The SEM followed closely, while the OLS model had the highest AIC (**Table 4**). Therefore, spatial dependencies play a role in explaining variations in COVID-19 CFR among Florida counties.

**Table 4:** Comparison of AIC values in OLS, SLM and SEM models for case rate, mortality rate and CFR.

| Outcome variables | OLS | SLM | SEM |
| --- | --- | --- | --- |
| Case Rate | 1350.8 | 1352.7 | 1352.8 |
| Mortality Rate | 836.8 | 838.2 | 837.8 |
| Case Fatality Rate | 32.3 | 29.9 | 31.2 |

## Discussion

Socioeconomically disadvantaged communities often bear a greater burden during epidemics [23,24], and disparities were evident during the COVID-19 pandemic [25,26]. In Florida, links between geographic factors and COVID-19 outcomes have previously been identified [27,28]. Here, we further explore these associations by comprehensively investigating the interplay between demographic, socioeconomic, and public health factors. Through our analysis, we observed significant temporospatial variations in county-level COVID-19 outcomes, which are explained in part by variation in socioeconomic and health-related risk factors as well as vaccination coverage.

While Florida’s epidemic from the onset of the pandemic until December 2022 followed the US trend, subsequent county-level analysis revealed several notable trends. Rural counties experienced higher case rates in the first and third waves than their urban counterparts. Moreover, during the pandemic’s second and fourth wave, rural counties experienced significantly higher mortality than urban counties as well as significantly higher cumulative CFRs, a finding consistent with other US-based studies [29,30].

Variation in CFR was also observed across the five waves, highlighting differences in epidemic trajectories between rural and urban counties, which corresponded with differences in vaccine uptake. Specifically, CFR remained elevated among rural counties during the fourth and fifth waves after the introduction of COVID-19 vaccines, which are highly efficacious in reducing mortality [31]. Mirroring the trend in CFR, all-cause mortality, which is robust to potential variation in COVID-19 mortality reporting among rural and urban counties, followed the same pattern. Together, these observations elucidate a broader and potentially longer-lasting impact of the pandemic. Detrimental collateral effects of the pandemic on the diagnosis and management of chronic health conditions have been described [32], and these effects often disproportionally impact rural communities.

Higher rates of SARS-CoV-2 transmission, limited access to tertiary healthcare, high prevalence of commodities, and decreased vaccine uptake may contribute to observed differences in COVID-19 outcomes. In general, rural populations tend to be un- or under-insured, older, more likely to live farther away from tertiary medical facilities and have underlying health conditions, which increase their likelihood for adverse outcomes [33]. Indeed, here we find that median income, adult obesity, smoking rates, and diabetes were associated with higher county-level mortality rates. Obesity, diabetes, and smoking are well-recognized risk factors for COVID-19 severity and mortality [34,35]. In univariate analysis, number of acute care and hospital beds and the proportion uninsured in a county were inversely associated with all COVID-19 outcomes, suggesting that access to care contributed to the observed disparities.

The relationship between race and ethnicity, socioeconomic status, and health is well established. Higher rates of crowding and increased household size are often observed among African American and Latinx communities [36], which in turn may result in more frequent person-to-person contact and increased SARS-CoV-2 transmission [37]. Furthermore, African Americans have higher prevalence of cardiovascular disease, obesity, diabetes and hypertension, which may contribute to COVID-19 severity [38,39]. Throughout the pandemic, race has been a consistent factor associated COVID-19 outcomes [40,41]. Here, we found a significant positive association between the percentage of Black persons in a county with case rate and CFR. Interestingly, while previous studies have reported increased COVID-19 cases and mortality rates among Hispanic/Latinx population in US [42,43], we did not observe a significant association between the percent Hispanic population in a county and increased morbidity and mortality in our study. However, we found that both the percentage of Black and Hispanic populations were significantly and positively associated with county-level immunization rates. As higher immunization rates were protective in multivariable models of mortality and CFR, this may partially explain the lack of association between Hispanic percentage and COVID-19 outcomes, while raising questions about why the percentage Black population remained associated with increased morbidity and mortality. Further research is required to better understand the factors of the observed variation in COVID-19 outcomes in Latinx and Black populations. Overall, pandemic preparedness should recognize the substantial impact of systemic flaws within the structural framework of American society, encompassing issues related to healthcare accessibility and the availability of community-level resources [44].

Our analysis also indicates that rural counties had slower vaccine uptake and significantly lower vaccination coverage than urban counties, which may have resulted in excess mortality during later pandemic waves. Indeed, county vaccination rates remained significantly associated with lower mortality rates when controlling for social and demographic variables. These findings align with recent studies conducted in the US [45,46]. Several factors may contribute to variation in vaccination coverage. First, healthcare access in rural counties remains a challenge, with residents of rural areas more likely to report decreased satisfaction with healthcare facilities than those in urban areas. Decrease healthcare access may have limited accessibility of COVID-19 vaccination [33]. Second, there are potential differences in knowledge and attitudes between urban and rural areas regarding the severity of COVID-19 and the implementation of mitigation strategies, which are influenced by different political ideologies and sociocultural identities [33]. Third, rural areas have historically higher vaccine hesitancy for routine vaccines, which likely contributed to reduced COVID-19 vaccination coverage. For example, during the pandemic, rural adults in the US were reported to be nearly three times more likely than urban adults to respond that they “definitely won’t” get vaccinated against COVID-19 [47]. Our multivariate analysis of county level predictors of vaccination coverage indicates that all these factors likely contributed to low vaccination rates (Supplementary Table S1). Further, our finding that increased county-level vaccination coverage was associated with decreased mortality rate and CFR indicates that a targeted approach to increasing vaccine confidence is needed to close the gap in vaccination coverage between rural and urban communities.

While this study provides valuable insights, several limitations warrant consideration. The use of cross-sectional data limits the ability to infer causal relationships. Additionally, other unmeasured factors, such as variation in county-level interventions, population mobility, and cultural practices, may also shape COVID-19 outcomes. In Florida in particular, there was significant variation in county-level mitigation strategies such as non-pharmaceutical interventions due to the absence of state-level policies.

Therefore, our analysis did not account for differences in mask mandates, stay-at-home orders, or school-based interventions. Last, an underlying assumption of our analysis is that case, death, and vaccine reporting was consistent among Florida counties. In particular, differences in testing capacity between rural and urban counties may have led to underreporting of cases among rural counties and a subsequent over-estimation of the CFR [48]. However, our sensitivity analysis suggests that our results are robust to modest differences in testing and case reporting, and our assessment of all-cause mortality in addition to COVID-19 mortality supports the observed disparities.

## Conclusion

In conclusion, this study contributes to our understanding of the dynamics of the COVID-19 pandemic in Florida and accentuates the vulnerability of rural communities to health crises. Spatial variation and associations between demographic, socioeconomic, and health factors emphasize the need for tailored and equitable public health strategies addressing infectious diseases and the broader determinants of health. The insights gained from this study have the potential to inform evidence-based interventions aimed at reducing health disparities and enhancing overall population resilience in the face of public health crises.

## Data Availability

All data produced in the present study are available upon reasonable request to the authors.

## Authorship contribution

**Sobur Ali**: Conceptualization, Data curation, Formal analysis, Investigation, Methodology, Validation, Visualization, Writing – original draft, Writing – review & editing **Taj Azarian:** Conceptualization, Project administration, Resources, Supervision, Writing – review & editing

---

**Supplementary Figure S1:**
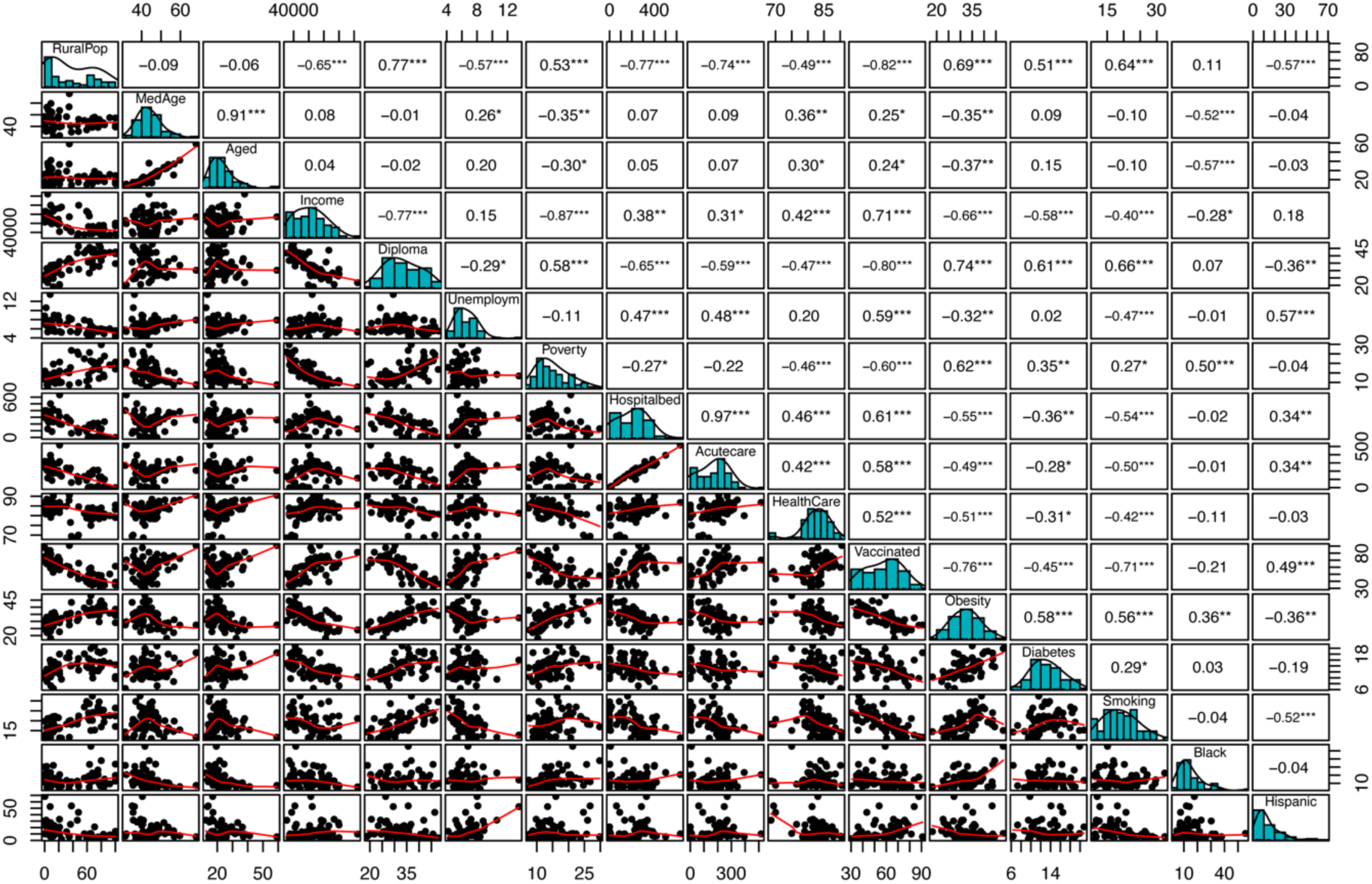
Correlation matrix of covariates. Spearman’s correlation coefficient was used to measure the correlation between predictor variables. The diagonal shows the distribution of each variable in the histogram. On the top are the values of the correlations with significance measures (asterisks), and the bivariate scatter plots are shown at the bottom. p-value: *** < 0.001; ** <0.01; * < 0.05.

**Supplementary Table S1.** Multivariable regression models of the association between socioeconomic characteristics and risk factors with the vaccination coverage for COVID-19. Vaccination coverage is measured per 100 people, and data was included from December 2020 to December 2022.

| Variables | Adjusted R square | Estimate | Std. Error | t value | Pr(>\|t\|) | VIF |
| --- | --- | --- | --- | --- | --- | --- |
| **Intercept** |  | -5.56 | 9.14 | -0.61 | 0.545 |  |
| Median Income |  | < 0.001 | < 0.001 | 6.57 | < 0.001*** | 1.91 |
| Unemployment Rate |  | 2.03 | 0.69 | 2.93 | 0.004** | 2.07 |
| Rural population (%) | 0.85 | -0.15 | 0.04 | -4.07 | < 0.001*** | 2.69 |
| % of black population |  | 0.32 | 0.09 | 3.55 | < 0.001*** | 1.37 |
| Aged over 65 years |  | 0.64 | 0.11 | 5.81 | < 0.001*** | 1.43 |
| % of Hispanic population |  | 0.23 | 0.07 | 3.10 | 0.003** | 1.79 |

